# Diagnostic competence and health worker knowledge of female genital schistosomiasis management in a rural Ghanaian district

**DOI:** 10.1101/2025.10.13.25337868

**Authors:** Courage Gbeze, Akosua Bonsu Karikari, Samuel Amoako Asirifi, Nana Boakye Alahaman, Seth Christopher Yaw Appiah, Clémence Essé-Diby, Gloria Ivy Mensah, Kennedy Kwasi Addo

## Abstract

**Background:** Schistosomiasis is a neglected tropical disease with a greater burden in Africa, including Ghana. Female Genital Schistosomiasis (FGS), a gynaecological manifestation of urogenital schistosomiasis, is often missed or misdiagnosed due to similarities with sexually transmitted infections and other gynaecological infections, with limited ease of health worker identification and diagnostic capability. This study assessed healthcare workers’ knowledge and diagnostic capacity for FGS in the Central Gonja District of Ghana.

**Methods:** A quantitative cross-sectional study sampled 237 healthcare workers from 19 facilities near the Black and White Volta rivers using a three-phase multistage sampling process. Data was collected via a self-administered Kobo Toolbox questionnaire, focusing on sociodemographic factors, whether facilities had functional screening tools for FGS and health workers’ capacity to diagnose and treat these conditions. Analysis was conducted in SPSS, employing descriptive statistics and Pearson’s chi-square tests to assess inferential associations between variables and health workers’ knowledge of FGS/schistosomiasis, which served as the main outcome variable.

**Results:** The study involved health workers with a mean age of 31.6 ± 4.18 years, of whom 52.3% were male. Knowledge gaps were significant: only 30% (71/237) demonstrated good understanding of schistosomiasis and merely 16.9% (40/237) showed adequate knowledge of FGS. Despite 91.6% recognition of schistosomiasis (‘Bilharzia’), knowledge of genital manifestations lagged severely (FGS: 26.8%, MGS (Male genital schistosomiasis): 18.1%). While demographic factors showed no association, experienced staff demonstrated better FGS knowledge (p = 0.003). Critical health system deficiencies emerged; 74% of facilities lacked laboratories, 90% lacked praziquantel and 100% lacked FGS diagnostic capacity. Even among clinicians, <43% knew standard FGS treatment and only 1/3 considered FGS in relevant diagnoses.

**Conclusion:** Healthcare workers lack FGS knowledge and diagnostic capacity, urgently requiring integration into reproductive health guidelines, training, and better resources for early detection and management

**Author Summary:** Female Genital Schistosomiasis is a neglected yet serious health condition affecting women, especially in sub-Saharan Africa. It often goes undiagnosed or is mistaken for sexually transmitted or other gynaecological infections due to overlapping symptoms and limited awareness among healthcare providers. This study explored how well healthcare workers in the Central Gonja District of Ghana understand and diagnose FGS. Through a survey of 237 healthcare workers from 19 health facilities located near the Black and White Volta rivers, we uncovered significant knowledge gaps; only a small number had a clear understanding of FGS and how to treat it. Most health facilities lacked the necessary resources, including diagnostic tools and essential medication like praziquantel. Our findings highlight an urgent need to train healthcare workers and integrate FGS into routine reproductive health services to improve detection and care for affected women.

## Introduction

Human schistosomiasis, or bilharzia, is a neglected tropical parasitic disease prevalent in communities lacking access to sufficient safe water and proper sanitation facilities (1). It is among the WHO’s twenty neglected tropical diseases (NTDs), a recognised disease of poverty with significant public health challenge in low and middle-income countries (2). Schistosomiasis has been reported in 78 countries worldwide, with almost 95% of the global cases being found in Africa (3). The disease causes debilitating consequences that is second only to malaria, exerting a significant socioeconomic impact as one of the most detrimental parasitic diseases (4). More than 779 million people are at risk of infection causing 280,000 deaths annually and a global burden of 3.3 million disability-adjusted life years (5–8)

The eggs of the parasite elicit an inflammatory immune response, leading to acute or chronic illness. While the disease is rarely fatal, its chronic form can cause significant health issues. *Schistosoma mansoni* and *S. intercalatum* infections are commonly associated with abdominal pain, bloody diarrhoea, high fever, hepatomegaly and hepatic fibrosis (9). Meanwhile, chronic infection with *S. haematobium* may result in haematuria, bladder irritation, bladder cancer, as well as painful and frequent urination (9). Prolonged infection with *S. haematobium* may lead to Female Genital Schistosomiasis (FGS) which is a gynaecological disease characterised by the presence of schistosome eggs and/or distinctive pathology in the female reproductive system. Eggs trapped in urogenital tissues trigger an inflammatory response, resulting in various lesions within the genital tract of affected women and girls (10,11).

FGS is a significant public health concern yet remains an overlooked and silent epidemic (12). Up to 150 million adolescent girls and women are estimated to be at risk of FGS (2,13), close to 16– 56 million women live with FGS, with majority resident in sub-Saharan Africa (14,15). The variability of these estimates’ points to the fact that this NTD is not well studied and often not prioritised by local, regional and global health policymakers (16,17). FGS is neither included in the training of healthcare professionals nor is it considered in the syndromic management of vaginal discharge, infertility or cervicovaginal lesions in endemic regions in Africa and may thus be mistakenly diagnosed as a sexually transmitted disease or cancer (18,19) in health centres, Community Health Planning and Services (CHPS) compounds, clinics, and even referral facilities like polyclinics and district hospitals. Despite sustained control efforts including mass praziquantel administration and school health education by the Ghana Health Service, schistosomiasis endemicity persists in Ghana’s freshwater communities where water-sanitation infrastructure is inadequate (20,21). The disease burden reflects systemic challenges in health worker capacity building, with insufficient post-curricular training on schistosomiasis pathology, especially FGS, resulting in underdiagnosis and mismanagement in endemic zones. This knowledge deficit contributes to low clinical awareness of FGS and its complications, even in endemic regions (21– 23).

This misdiagnosis is further compounded when patients are treated for sexually transmitted infections (STIs), being administered other medications such as antibiotics or antifungal medications, which have minimal to no effect on FGS (22) instead of praziquantel, the drug of choice for treating all forms of schistosomiasis. In addition to physical complications, FGS could lead to outcomes such as miscarriage and infertility, which may significantly impact women’s mental health and social status (24). In addition to these, misdiagnosis often results in repeated visits to health facilities, increasing the strain on both the patient and the healthcare system (22).This study sought to investigate the knowledge level and capacity of healthcare workers to diagnose and manage FGS in an endemic district of the Savannah region of Ghana.

## Method

### Ethical considerations

Ethical approval was obtained from the Institutional Review Board (IRB) of the University for Development Studies (UDS), Tamale-Ghana with approval number UDS/RB/030/24. Permission was sought from the district director of health services as well as heads of the various health institutions. All health workers who participated in the study provided consent. Participant confidentiality was ensured such that no personal identifiers were cited in the questionnaire and no personal data was recorded since the questionnaire was accessed through an online link.

### Study area

This study was conducted in the central Gonja district of the Savannah region in Ghana. The district lies between longitudes 1°5’W and 2°58’W and latitudes 8°32’N and 10°2’N. The 2021 national population census reports a total of 116,873 inhabitants residing in the district (source: GSS 2021 PHC). The district has two major rivers, that is the Black and the White Volta, which coalesce/confluence at Kikale. The study area was purposively chosen because of the freshwater source, which is a favourable breeding ground for the intermediate host snail of schistosomes. The inhabitants of this district are mostly farmers, with majority of them either living closer to the rivers or are fishermen and the women are dominantly fishmongers. The district is mostly rural, with the capital, Buipe, being peri-urban. According to the Country Health Information Platform (CHIP), there is moderate prevalence of schistosomiasis in the district, with this value ranging between 10-49% (25).

### Study design and sampling

The study employed a quantitative approach using a cross-sectional design. A multi-stage sampling strategy was implemented. In the first stage, purposive sampling was used to select health facilities located near the two major rivers, Black and White Volta, as well as all referral polyclinics and the district hospital in the Central Gonja District. Initially, 24 health facilities were targeted, but only 19 consented to participate in the study. In the second stage, a stratified sampling technique was applied. Each participating facility was stratified into six professional categories based on staff roles: nursing, midwifery, diagnostics, prescribers, pharmacy, and public health. At the third stage, a simple random sampling method was used to select eligible healthcare workers from each stratum. Eligibility was based on having worked in the respective unit for at least six months. All selected participants were enrolled in the study after providing informed consent. A self-administered questionnaire was used to collect data. The data collection was done between June 2024 and October 2024. The questionnaire subdivided into seven sections: Demographic Data, Facility Data, KAP data (knowledge indicators or familiarity with the signs and symptoms, modes of transmission, control methods and risk factors), Capacity of Health Facilities in Treating FGS, Laboratory Capacity to Diagnose schistosomiasis & FGS, Clinical Care and Perception of Health Workers on schistosomiasis & FGS

### Sample size estimation

Study participants were health workers from various health facilities in the district. A total number of 237 responses were received at the end of data collection out of 354 questionnaires issued.

Using the Slovin formula 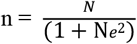, a sample size of 226 was calculated.

Where n = Sample size

N= Population size = 354

e = Acceptable margin of error = 0.04

### Data collection

In each unit within each health facility, all staff members were considered in the sample frame and as such, each person stood an equal chance of being recruited. Various units within the health facilities were visited, and informed consent was obtained from eligible individuals. All those who met the inclusion criteria and consented to participate were enrolled until the conclusion of the data collection period. An open-access data collection tool, Kobo Collect (26) was shared with all the staff in each unit using a generated online form for them to respond to the questionnaire. The questionnaire was piloted at St. Anne Hospital, Damongo, among a small group of respondents to ensure clarity and relevance. Feedback from the pilot informed minor revisions before the main data collection.

### Data analysis

The collected data in Kobo Collect was downloaded as an Excel file, cleaned and uploaded into Statistical Package for Social Sciences (SPSS) version 20 for analysis. Summarised descriptive statistics (frequency values and percentages) and crosstabulations were obtained for the Knowledge variables and the respondents’ demographic profile. Contingency tables were created, and Pearson’s chi-square test (χ2) was used to analyse nominal variables and to examine the association of selected variables with the two main outcome variables of interest; knowledge of schistosomiasis and knowledge of female genital schistosomiasis and considered statistically significant at a P < 0.05.

## Results

### Demographic Characteristics

The ages of the 237 respondents ranged between 23 and 49 years, with a median of 31 years and a mean of 31.60 ± 4.178 years. There were slightly more males, 124 (52.3%), than female 113(47.7%) health workers enrolled, with the working years spanning from a few 2(0.8%) working under two years to a maximum of 19 years, while the mean working years for the health workers was 3.97 ± 3.134. Only the district hospital was a secondary healthcare facility. Of the total number of persons sampled, the majority 88 (37.1%) were recruited from the district hospital followed by those working at policlinics 79 (33.3%) with the bulk of healthcare workers sampled being Enrolled Nurses (74; 31.2%) followed by Nurses (60; 25.3%) and Midwives (24; 10.1%). Two of the health workers held master’s degrees, 44 persons with bachelor’s degrees and the least qualification of a certificate was reported among 107 health workers.

### Awareness and Knowledge of Schistosomiasis and FGS

Figure 2a highlights the generally low knowledge levels of schistosomiasis and FGS among health workers in the Central Gonja District. Of the 237 respondents, 71(30.0%) demonstrated good knowledge of schistosomiasis, while the majority, 166 (70.0%), exhibited poor knowledge. Similarly, knowledge of FGS was limited, with 40 (16.9%) showing good understanding, whereas 197 (83.1%) had poor knowledge. Figure 2b shows the number of respondents who knew and had heard of the various forms of schistosomiasis. As it is commonly known, Bilharzia was selected by 91.6% of the respondents while 67.93% selected knowing schistosomiasis. We enquired if respondents knew schistosomiasis to be the same as bilharzia, and 70.5% affirmed. About 32.9% and 29.1% knew of Urogenital Schistosomiasis and Intestinal Schistosomiasis, respectively. Only 26.6% and 18.1% knew about FGS and MGS, respectively.

**Fig. 1.**
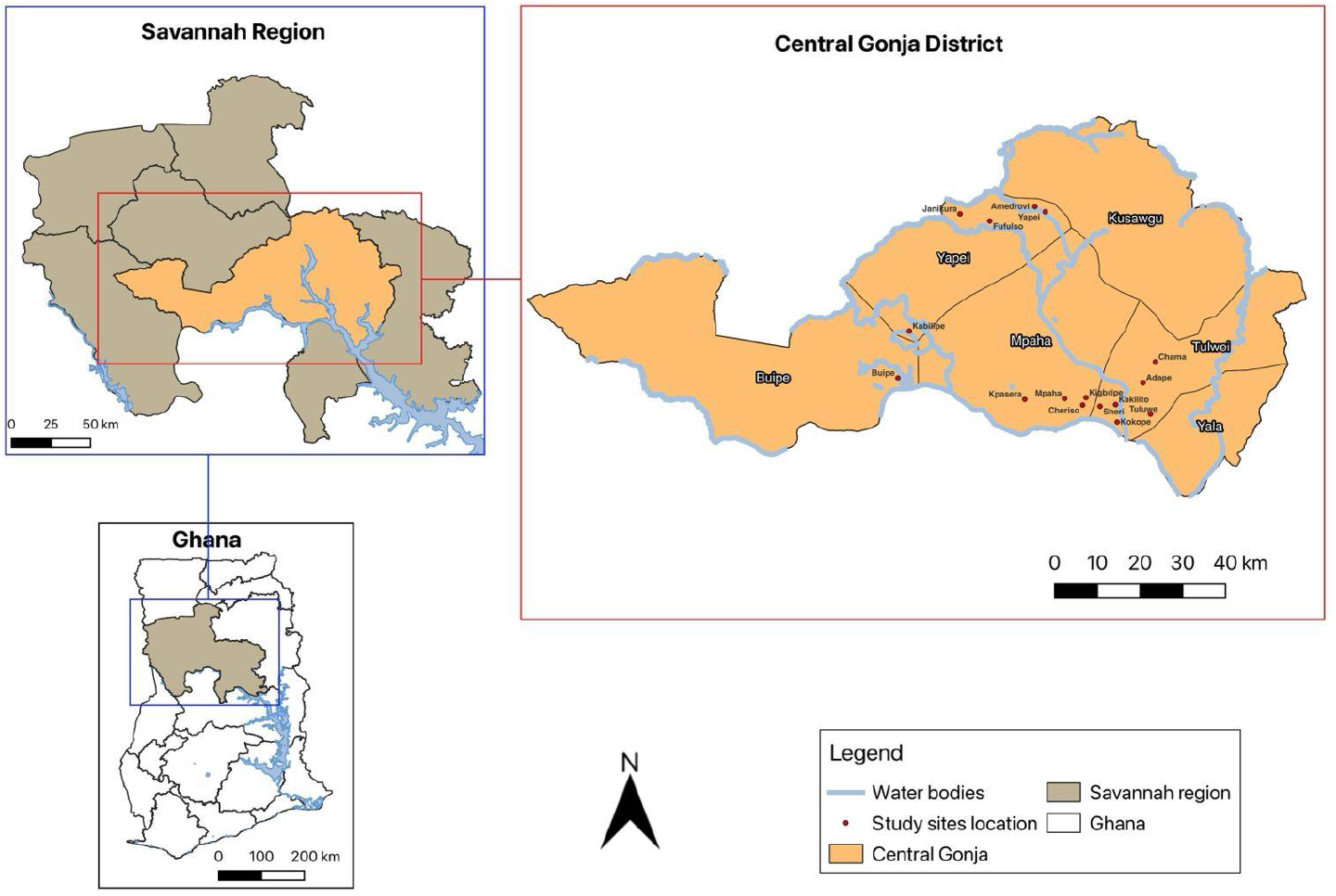
Picture showing the study sites in the central Gonja district of the savannah region in Ghana.

**Fig 2:**
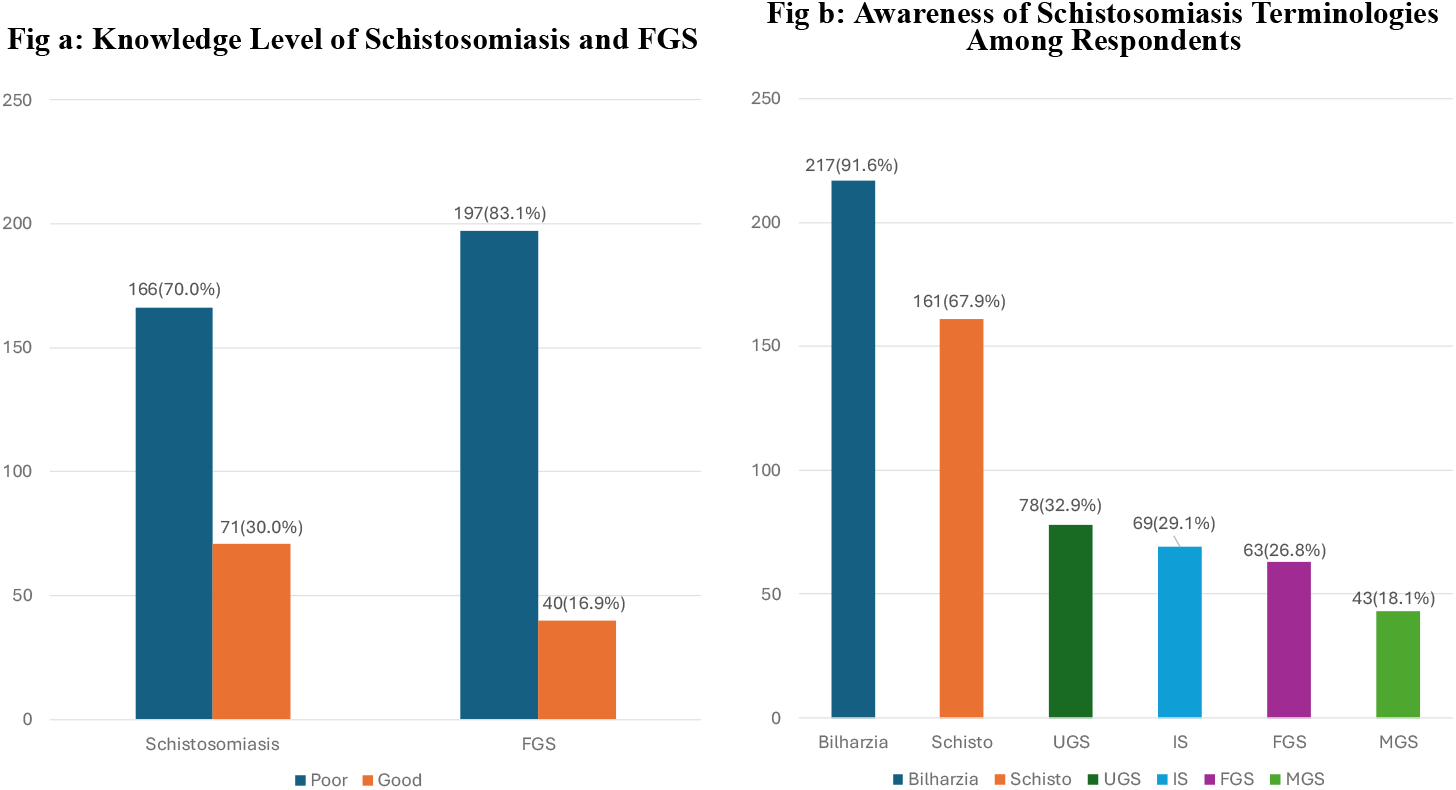
Awareness and Knowledge level of Schistosomiasis and FGS. *\*FGS - Female Genital Schistosomiasis, Schisto – Schistosomiasis, UGS - Urogenital Schistosomiasis, IS – Intestinal Schistosomiasis, MGS – Male Genital Schistosomiasis.*

For enhancing awareness of FGS among health workers, respondents suggested multiple strategies. Popular approaches were to conduct “workshops” and “in-service training” sessions to “educate and enlighten staff” on FGS, its effects and management. Additionally, respondents emphasised “health education” through “public education,” such as community “durbars” and media engagement, along with resources like “fliers”, “clinical presentations” and “posters” to reinforce knowledge and visibility. They also suggested “curriculum adjustments” in training institutions and interprofessional meetings as essential measures to foster a comprehensive understanding and proactive engagement with FGS in healthcare settings.

### Knowledge Level of Healthcare Workers on Schistosomiasis and Female Genital Schistosomiasis

Overall, the mean score for both schistosomiasis and FGS knowledge levels were 10.02 ± 7.315 and 4.02 ± 6.509, indicating a generally poor knowledge of respondents on the subject matters, with a slightly better understanding of schistosomiasis. The large standard deviations for both scores suggest wide variation in knowledge levels among participants, with a few having considerably higher scores.

**Table 2** presents the distribution of knowledge levels among healthcare workers in the Central Gonja District concerning schistosomiasis and FGS. The analysis considers demographic characteristics, educational background, job title and professional experience to assess variations in knowledge levels. Among the 237 respondents, males (52.3%) had a slightly higher good knowledge on schistosomiasis (33.1%) compared to females (26.5%), though the difference was not statistically significant (χ^2^ = 1.196, p = 0.274). Similarly, for FGS, 19.4% of males exhibited good knowledge compared to 14.2% of females, with no significant difference (χ^2^ = 1.138, p = 0.286). Across educational levels, knowledge of schistosomiasis and FGS did not show a significant association. Healthcare workers were grouped based on job roles, including diagnostic staff, midwives, nurses, pharmacy staff, prescribers and public health staff. While there were variations in knowledge levels, the associations were not statistically significant for schistosomiasis (χ^2^ = 10.034, p = 0.074) and FGS (χ^2^ = 5.4, p = 0.369). Notably, pharmacy staff had the highest proportion of good schistosomiasis knowledge (70.0%), while midwives had the lowest (16.7%). For FGS, diagnostic staff (31.3%) and prescribers (30.0%) demonstrated better knowledge compared to nurses (15.7%) and midwives (8.3%). Tenure categories revealed significant differences in knowledge levels for FGS (χ^2^ = 15.673, p = 0.003) but not for schistosomiasis (χ^2^ = 8.42, p = 0.077). Highly experienced healthcare workers (19+ years) exhibited the poorest knowledge of both schistosomiasis and FGS, whereas those in the experienced career level (14–18 years) had the highest good knowledge proportions (100%) for both diseases. Entry-level (0–3 years) workers had the lowest FGS knowledge, with only 14.9% demonstrating good knowledge.

**Table 1:** Demographic Characteristics.

| Characteristics | Description | Frequency | Percent (%) |
| --- | --- | --- | --- |
| Age | Mean ± Std. Deviation = 31.6 ± 4.178 | Median=31 | Mode=29 |
| Gender | Female | 113 | 47.7 |
|  | Male | 124 | 52.3 |
| Educational Level | Certificate | 107 | 45.1 |
|  | Degree | 44 | 18.6 |
|  | Diploma | 84 | 35.4 |
|  | Masters | 2 | 0.8 |
| Job Title Grouping | Diagnostic Staff | 16 | 6.8 |
|  | Midwives | 24 | 10.1 |
|  | Nurses | 134 | 56.5 |
|  | Pharmacy Staff | 10 | 4.2 |
|  | Prescribers | 10 | 4.2 |
|  | Public Health Staff | 43 | 18.1 |
| Tenure Categories | Entry Level (0 - 3 yrs) | 154 | 65 |
|  | Early Career Level (4 - 8 yrs) | 58 | 24.5 |
|  | Mid-Career Level (9 - 13 yrs) | 21 | 8.9 |
|  | Experienced Career Level(14-18yr) | 3 | 1.3 |
|  | Highly Experienced Career Level (19+ yrs) | 1 | 0.4 |
| Facility | Adape CHPS | 3 | 1.3 |
|  | Amedrovi CHPS | 1 | 0.4 |
|  | Bonyase CHPS | 2 | 0.8 |
|  | Buipe Polyclinic | 62 | 26.2 |
|  | Central Gonja District Hospital | 88 | 37.1 |
|  | Chama CHPS | 3 | 1.3 |
|  | Cheriso CHPS | 1 | 0.4 |
|  | Fufulso Health Center | 14 | 5.9 |
|  | Holistic Medicare | 1 | 0.4 |
|  | Janikura CHPS | 2 | 0.8 |
|  | Kigbripe CHPS | 1 | 0.4 |
|  | Kokoje CHPS | 2 | 0.8 |
|  | Kpasera CHPS | 3 | 1.3 |
|  | Mpaha Health Centre | 25 | 10.5 |
|  | Sheri CHPS | 2 | 0.8 |
|  | Tuluwe CHPS | 3 | 1.3 |
|  | Yapei Polyclinic | 18 | 7.6 |
|  | Yapei Quarters CHPS | 4 | 1.7 |
|  | Yapei Yipala CHPS | 2 | 0.8 |
| **Respondents from Facility Type** | CHPS | 26 | 11 |
|  | DISTRICT HOSPITAL | 90 | 38 |
|  | Health Centre | 43 | 18.1 |
|  | POLYCLINIC | 77 | 32.5 |
|  | Private Health Facility | 1 | 0.4 |
| **Facility Type (N=19)** | CHPS | 13 | 68.4 |
|  | Polyclinic | 2 | 10.5 |
|  | District Hospital | 1 | 5.3 |
|  | Private Health Facility | 1 | 5.3 |
|  | Health Centre | 2 | 10.5 |

**Table 2:**
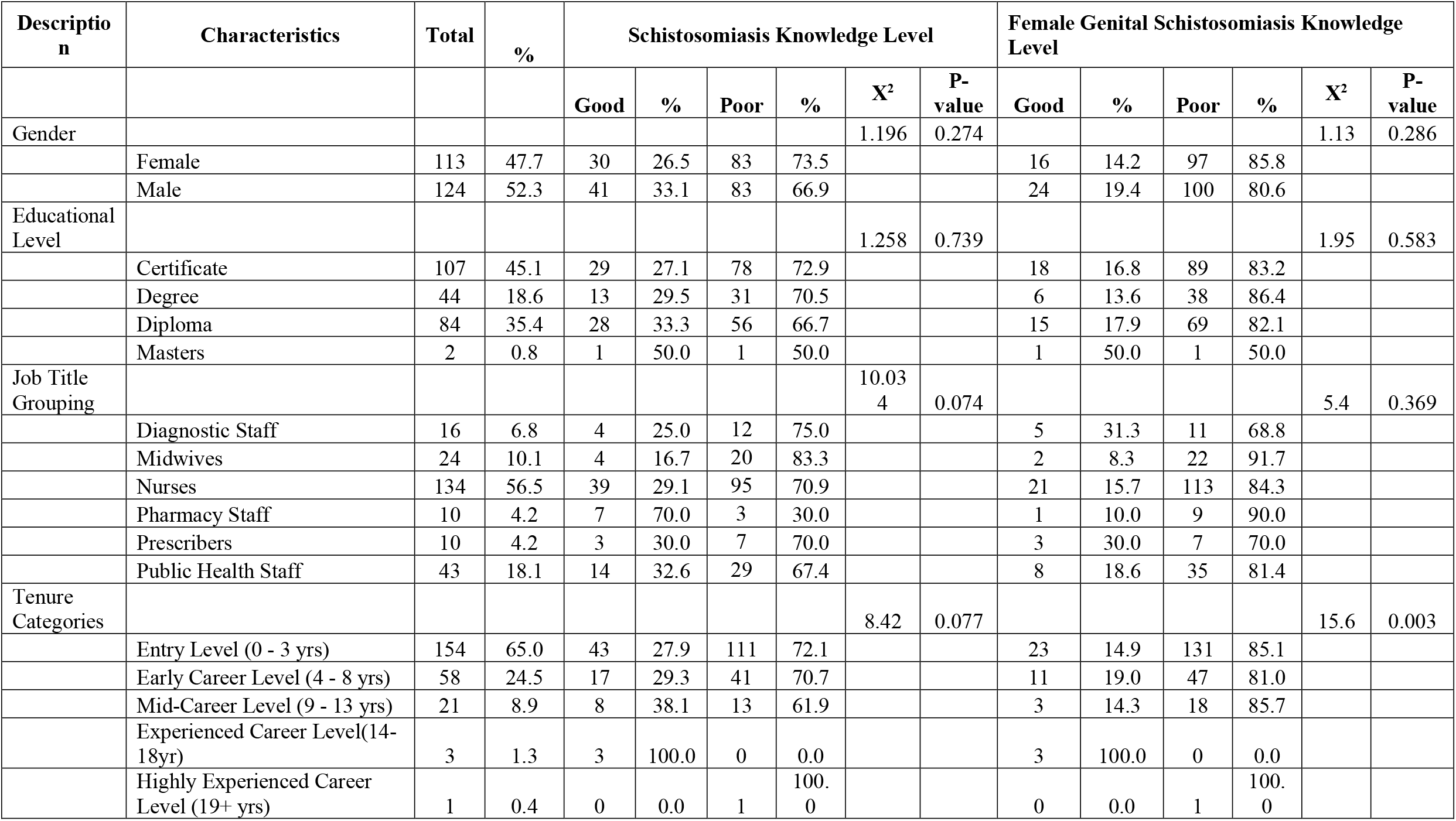
Association of Schistosomiasis and FGS Knowledge and Demographic Characteristics.

| Description | Characteristics | Total | % | Schistosomiasis Knowledge Level |  |  |  |  |  | Female Genital Schistosomiasis Knowledge Level |  |  |  |  |  |
| --- | --- | --- | --- | --- | --- | --- | --- | --- | --- | --- | --- | --- | --- | --- | --- |
|  |  |  |  | Good | % | Poor | % | X<sup>2</sup> | P-value | Good | % | Poor | % | X<sup>2</sup> | P-value |
| Gender |  |  |  |  |  |  |  | 1.196 | 0.274 |  |  |  |  | 1.13 | 0.286 |
|  | Female | 113 | 47.7 | 30 | 26.5 | 83 | 73.5 |  |  | 16 | 14.2 | 97 | 85.8 |  |  |
|  | Male | 124 | 52.3 | 41 | 33.1 | 83 | 66.9 |  |  | 24 | 19.4 | 100 | 80.6 |  |  |
| Educational Level |  |  |  |  |  |  |  | 1.258 | 0.739 |  |  |  |  | 1.95 | 0.583 |
|  | Certificate | 107 | 45.1 | 29 | 27.1 | 78 | 72.9 |  |  | 18 | 16.8 | 89 | 83.2 |  |  |
|  | Degree | 44 | 18.6 | 13 | 29.5 | 31 | 70.5 |  |  | 6 | 13.6 | 38 | 86.4 |  |  |
|  | Diploma | 84 | 35.4 | 28 | 33.3 | 56 | 66.7 |  |  | 15 | 17.9 | 69 | 82.1 |  |  |
|  | Masters | 2 | 0.8 | 1 | 50.0 | 1 | 50.0 |  |  | 1 | 50.0 | 1 | 50.0 |  |  |
| Job Title Grouping |  |  |  |  |  |  |  | 10.034 | 0.074 |  |  |  |  | 5.4 | 0.369 |
|  | Diagnostic Staff | 16 | 6.8 | 4 | 25.0 | 12 | 75.0 |  |  | 5 | 31.3 | 11 | 68.8 |  |  |
|  | Midwives | 24 | 10.1 | 4 | 16.7 | 20 | 83.3 |  |  | 2 | 8.3 | 22 | 91.7 |  |  |
|  | Nurses | 134 | 56.5 | 39 | 29.1 | 95 | 70.9 |  |  | 21 | 15.7 | 113 | 84.3 |  |  |
|  | Pharmacy Staff | 10 | 4.2 | 7 | 70.0 | 3 | 30.0 |  |  | 1 | 10.0 | 9 | 90.0 |  |  |
|  | Prescribers | 10 | 4.2 | 3 | 30.0 | 7 | 70.0 |  |  | 3 | 30.0 | 7 | 70.0 |  |  |
|  | Public Health Staff | 43 | 18.1 | 14 | 32.6 | 29 | 67.4 |  |  | 8 | 18.6 | 35 | 81.4 |  |  |
| Tenure Categories |  |  |  |  |  |  |  | 8.42 | 0.077 |  |  |  |  | 15.6 | 0.003 |
|  | Entry Level (0 - 3 yrs) | 154 | 65.0 | 43 | 27.9 | 111 | 72.1 |  |  | 23 | 14.9 | 131 | 85.1 |  |  |
|  | Early Career Level (4 - 8 yrs) | 58 | 24.5 | 17 | 29.3 | 41 | 70.7 |  |  | 11 | 19.0 | 47 | 81.0 |  |  |
|  | Mid-Career Level (9 - 13 yrs) | 21 | 8.9 | 8 | 38.1 | 13 | 61.9 |  |  | 3 | 14.3 | 18 | 85.7 |  |  |
|  | Experienced Career Level(14-18yr) | 3 | 1.3 | 3 | 100.0 | 0 | 0.0 |  |  | 3 | 100.0 | 0 | 0.0 |  |  |
|  | Highly Experienced Career Level (19+ yrs) | 1 | 0.4 | 0 | 0.0 | 1 | 100.0 |  |  | 0 | 0.0 | 1 | 100.0 |  |  |

Overall, knowledge of both schistosomiasis and FGS among healthcare workers was low, with most respondents demonstrating poor knowledge. Gender, educational level, and job title did not show significant associations with knowledge levels, while professional experience significantly influenced FGS knowledge.

### Facility’s Capacity for Diagnosing and Treating FGS

The data in **Table 3** shows significant disparities in the availability of laboratories and medications for treating schistosomiasis across healthcare facilities. The table highlights that most higher-level facilities, such as Buipe Polyclinic and Central Gonja District Hospital, reported laboratory presence, while smaller facilities like CHPS compounds (e.g., Adape, Amedrovi, and Bonyase) and health centres (e.g., Fufulso Health Centre) lacked laboratory services. Out of 19 facilities surveyed, only 5 reported having laboratories, with 14 indicating none. Laboratories in larger facilities are equipped with basic tools like microscopes but lack advanced diagnostic equipment, limiting their capacity for FGS diagnosis.

**Table 3:**
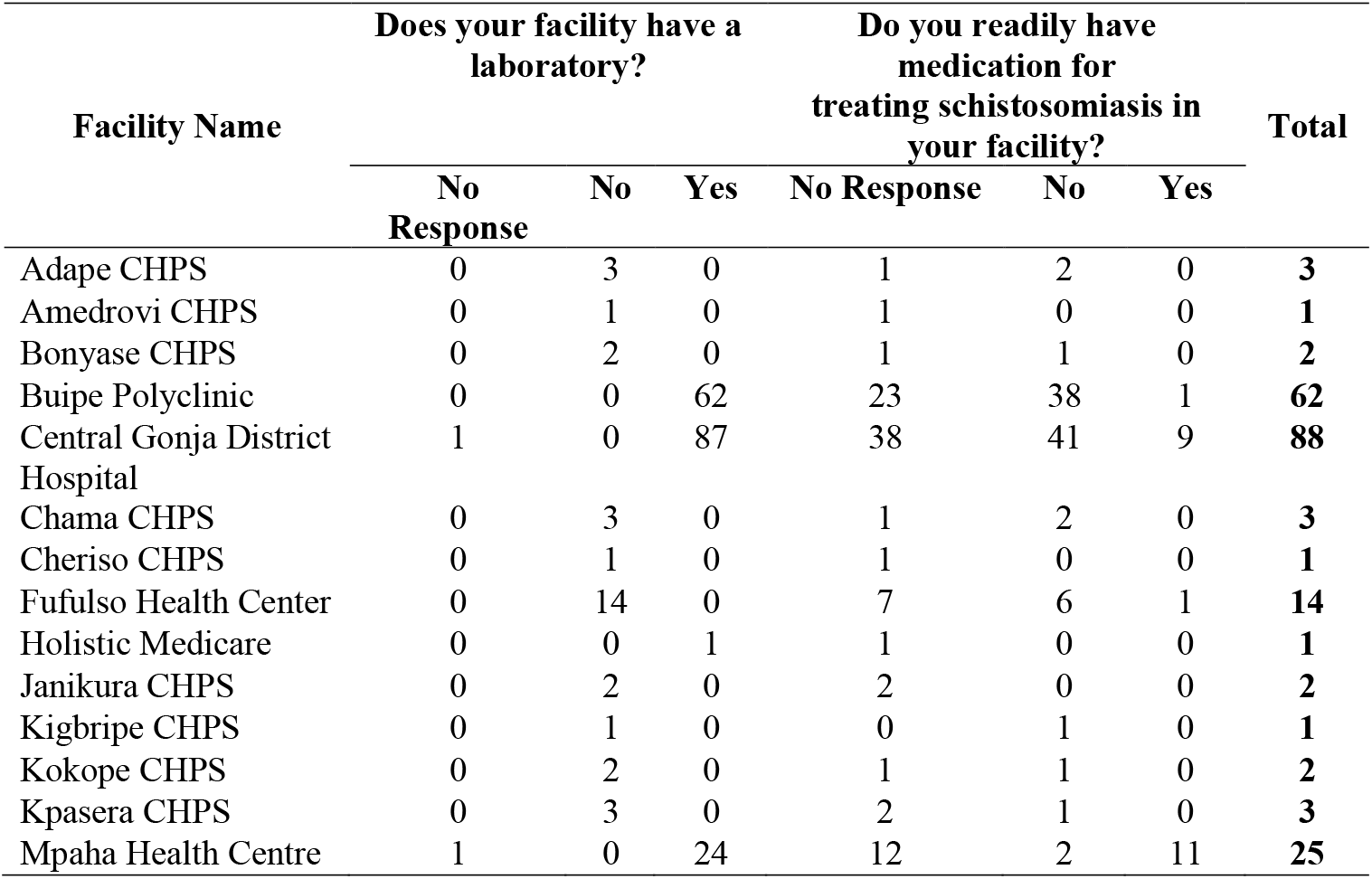
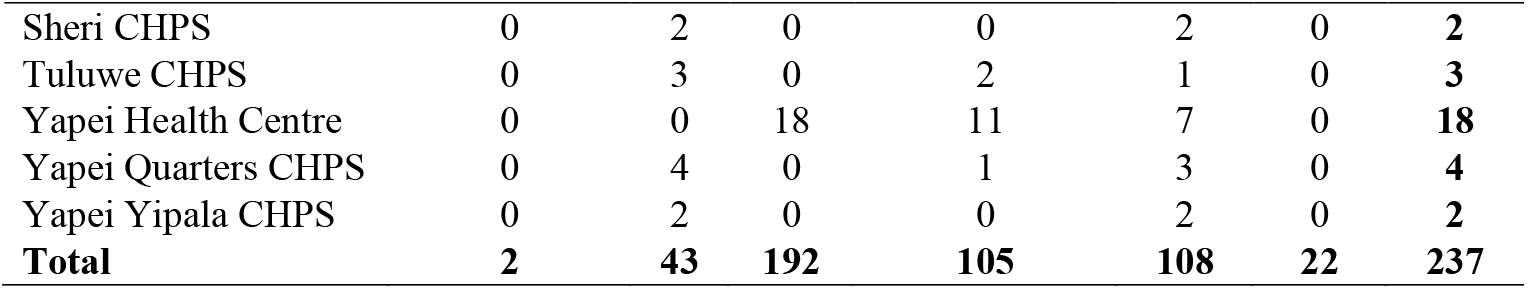
Availability of aboratories and Schistosomiasis Medication by Facility.

There is variability in the availability of medications for treating schistosomiasis. Larger facilities like Central Gonja District Hospital and Buipe Polyclinic reported the highest availability, with Central Gonja District Hospital having the most “Yes” responses, 10.2% (9/88). In contrast, smaller facilities, such as CHPS compounds and some health centres, either lack schistosomiasis medication or have staff not directly involved in patient treatment, as indicated by no responses. Only 10.1% (24/237) of responses confirmed the availability of schistosomiasis medication, underscoring limited access in smaller community-based facilities and highlighting disparities in resource distribution between facility levels.

Notably, none of the healthcare facilities surveyed had a colposcope for vaginal examination. While 23 respondents initially indicated that their facility had one, follow-up investigations revealed that these responses were likely due to misidentification. Additionally, 30 respondents were unsure about their facility’s colposcope availability, particularly in larger facilities like Central Gonja District Hospital and Buipe Polyclinic.

### Association between job categories and diagnostic capacities, knowledge and skills for identifying and managing Female Genital Schistosomiasis

**Table 4** summarises responses from 96 clinical staff directly involved in patient care regarding their knowledge and practices related to FGS. Among the respondents, the correct treatment was Praziquantel (PZQ), cited by (41 (42.7%) individuals, followed by 38(39.6%) citing antibiotics such as Ceftriaxone (CTX) and antifungal treatments like Fluconazole (FLZ). Notably, 17.7% (17) of the respondents indicated they “don’t know” how to treat FGS, highlighting a knowledge gap. Regarding consideration of FGS when female clients present with symptoms of STIs/STDs, only 33.3% (32) of respondents reported doing so, while 43.8% (42) did not and 22.9% (22) did not respond. A similar trend was observed in differentiating FGS from STIs/STDs and other vaginal infections, where 51.0% (49) respondents stated they could not make this distinction, 26.0% (25) could and 22.9% (22) did not respond. Furthermore, when asked if FGS is considered during differential diagnosis for vaginal infections resembling STIs, 46.9% (45) indicated they do not consider it, 30.2% (29) reported they do, and 22.9% (22) did not respond. Majority of respondents were nurses and midwives and accounted for the largest groups in categories reflecting limited knowledge and awareness, underscoring the need for enhanced training and education on FGS diagnosis and treatment, especially among frontline healthcare providers.

**Table 4:** Treatment and Consideration of Female Genital Schistosomiasis by Job Title/Occupation.

| Job Title/Occupation | How do you treat FGS? |  |  |  |  | Do you consider FGS when female clients present with symptoms of STIs/STDs? |  |  | Are you able to differentiate FGS from STIs/STDs and other vaginal infections? |  |  | Do you consider FGS during differential diagnosis when clients present with vaginal infections similar to STIs? |  |  | Total |
| --- | --- | --- | --- | --- | --- | --- | --- | --- | --- | --- | --- | --- | --- | --- | --- |
|  | No Response | PZQ | CTX | FLZ | Don't know | No Response | No | Yes | No Response | No | Yes | No Response | No | Yes |  |
| Facility In-Charge | 0 | 1 | 0 | 1 | 1 | 0 | 0 | 3 | 0 | 3 | 0 | 0 | 0 | 3 | 3 |
| Medical Doctor | 0 | 2 | 0 | 0 | 0 | 0 | 0 | 2 | 0 | 1 | 1 | 0 | 0 | 2 | 2 |
| Midwife | 3 | 9 | 2 | 2 | 8 | 3 | 17 | 4 | 3 | 17 | 4 | 3 | 16 | 5 | 24 |
| Nurse | 18 | 22 | 6 | 5 | 8 | 18 | 21 | 20 | 18 | 24 | 17 | 18 | 23 | 18 | 59 |
| Nurse Practitioner | 1 | 3 | 0 | 0 | 0 | 1 | 1 | 2 | 1 | 0 | 3 | 1 | 2 | 1 | 4 |
| Physician Assistant | 0 | 4 | 0 | 0 | 0 | 0 | 3 | 1 | 0 | 4 | 0 | 0 | 4 | 0 | 4 |
| Total | 22 | 41 | 8 | 8 | 17 | 22 | 42 | 32 | 22 | 49 | 25 | 22 | 45 | 29 | 96 |

\* PZQ = Praziquantel (Anthelmintic) CTX = Ceftriaxone (Antibiotic) FLZ = Fluconazole (Antifungal)

**Table 5:** Perceptions of Respondents on Genital Infections and Schistosomiasis.

| Job Title/Occupation | Do you consider clients to be promiscuous when they present with genital infections? |  |  | Do you consider Schistosomiasis to be a public health concern? |  |  | Do you consider FGS to be a threat to women's reproductive health? |  |  | Do you see the need for awareness creation and education on FGS? |  |  | Total |
| --- | --- | --- | --- | --- | --- | --- | --- | --- | --- | --- | --- | --- | --- |
|  | Did Not Respond | No | Yes | Did Not Respond | No | Yes | Did Not Respond | No | Yes | Did Not Respond | No | Yes |  |
| Community Health Officer | 4 | 0 | 0 | 0 | 0 | 4 | 0 | 0 | 4 | 0 | 0 | 4 | 4 |
| Community Health Nurse | 18 | 2 | 1 | 0 | 0 | 21 | 0 | 0 | 21 | 0 | 0 | 21 | 21 |
| Community Health Officer | 0 | 0 | 1 | 0 | 0 | 1 | 0 | 0 | 1 | 0 | 0 | 1 | 1 |
| Disease Control Officer | 3 | 0 | 0 | 0 | 0 | 3 | 0 | 0 | 3 | 0 | 0 | 3 | 3 |
| Dispensing Assistant | 1 | 0 | 0 | 0 | 0 | 1 | 0 | 0 | 1 | 0 | 0 | 1 | 1 |
| Enrolled Nurse | 67 | 2 | 3 | 0 | 15 | 57 | 0 | 1 | 71 | 0 | 1 | 71 | 72 |
| Facility In-Charge | 0 | 1 | 2 | 0 | 1 | 2 | 0 | 0 | 3 | 0 | 0 | 3 | 3 |
| Field Technician | 5 | 1 | 0 | 0 | 0 | 6 | 0 | 1 | 5 | 0 | 0 | 6 | 6 |
| Health Information Officer | 4 | 0 | 0 | 1 | 0 | 3 | 1 | 0 | 3 | 1 | 0 | 3 | 4 |
| Health Promotion Officer | 2 | 0 | 0 | 0 | 0 | 2 | 0 | 0 | 2 | 0 | 0 | 2 | 2 |
| Medical Doctor | 0 | 2 | 0 | 0 | 0 | 2 | 0 | 0 | 2 | 0 | 0 | 2 | 2 |
| Medical Laboratory Scientist | 1 | 2 | 3 | 0 | 1 | 5 | 0 | 0 | 6 | 0 | 0 | 6 | 6 |
| Medical Laboratory Technician | 0 | 6 | 2 | 0 | 0 | 8 | 0 | 0 | 8 | 0 | 0 | 8 | 8 |
| Midwife | 0 | 18 | 6 | 0 | 1 | 23 | 0 | 1 | 23 | 0 | 1 | 23 | 24 |
| Nurse | 0 | 43 | 16 | 0 | 4 | 55 | 0 | 2 | 57 | 0 | 3 | 56 | 59 |
| Nurse Practitioner | 0 | 4 | 0 | 0 | 0 | 4 | 0 | 0 | 4 | 0 | 0 | 4 | 4 |
| Pharmacist | 2 | 0 | 0 | 0 | 0 | 2 | 0 | 0 | 2 | 0 | 0 | 2 | 2 |
| Pharmacy Technician | 7 | 0 | 0 | 0 | 1 | 6 | 0 | 0 | 7 | 0 | 0 | 7 | 7 |
| Physician Assistant | 3 | 1 | 0 | 0 | 1 | 3 | 0 | 0 | 4 | 0 | 0 | 4 | 4 |
| Public Health Nurse | 0 | 0 | 1 | 0 | 0 | 1 | 0 | 0 | 1 | 0 | 0 | 1 | 1 |
| Public Health Officer | 1 | 0 | 0 | 0 | 0 | 1 | 0 | 0 | 1 | 0 | 0 | 1 | 1 |
| Sonographer | 1 | 0 | 0 | 0 | 0 | 1 | 0 | 0 | 1 | 0 | 0 | 1 | 1 |
| X-ray technician | 1 | 0 | 0 | 0 | 0 | 1 | 0 | 0 | 1 | 0 | 0 | 1 | 1 |
| **Total** | **120** | **82** | **35** | **1** | **24** | **212** | **1** | **5** | **231** | **1** | **5** | **231** | **237** |

### Perceptions of Respondents on Genital Infections and Schistosomiasis

Out of 237 respondents, 207 were clinical staff who responded to the question on whether they consider clients to be promiscuous when they present with genital infections, a majority (82 respondents) answered “No,” while 35 respondents indicated “Yes,” and 91 did not respond plus 29 that were not clinical staff. The data revealed that “Yes” responses were more common among nurses (16), midwives (6), and enrolled nurses (3). Notably, only a few medical doctors and laboratory scientists held this perception, suggesting that attitudes toward clients with genital infections may vary by job role.

Majority of respondents (212) consider schistosomiasis as a public health concern, reflecting widespread awareness of its implications among healthcare workers. Only 24 respondents disagreed. Similarly, 231 respondents recognised FGS as a threat to women’s reproductive health, with just 5 respondents not viewing it as a concern. Additionally, 231 respondents acknowledged the need for awareness creation and education on FGS, indicating strong support for initiatives to enhance knowledge and interventions around FGS. These findings demonstrate a shared understanding among healthcare providers of the significance of schistosomiasis and FGS, emphasising the importance of continued advocacy, education, and resource allocation to address these public health challenges effectively.

## Discussion

This is a pioneer prevalence study on schistosomiasis conducted in this district, including UGS, IS and FGS. This study assessed the knowledge level of health workers in the central Gonja district on female genital schistosomiasis following an ongoing prevalence study. Female Genital Schistosomiasis is a neglected tropical disease caused by chronic infection with *Schistosoma* species, primarily *Schistosoma haematobium*. It is a significant public health concern in endemic regions, particularly in Sub-Saharan Africa, where it disproportionately affects women of reproductive age. FGS results from the deposition of schistosome eggs in the genital tract, leading to inflammation, ulceration and scarring (27,28). These manifestations are often misdiagnosed as sexually transmitted infections or other gynaecological conditions due to overlapping clinical features and limited awareness among healthcare professionals. The clinical and socioeconomic impacts of FGS are profound. In addition to its debilitating symptoms, FGS increases susceptibility to HIV infection, infertility and poor pregnancy outcomes, thereby posing a significant burden on affected communities (28–30). Despite these implications, the knowledge and recognition of FGS among healthcare professionals remain insufficient, contributing to misdiagnoses.

. Health workers had poor knowledge of schistosomiasis and FGS. However, the mean score for schistosomiasis was better than FGS. Bilharzia was the most common name referred to by respondents as schistosomiasis with a lesser percentage of them not knowing or unsure. More respondents knew about urinary schistosomiasis as compared to FGS and MGS which aligns with other similar studies (1,31–35). This was posited to be the case by Wambui et al. (1) because blood in urine is the hallmark symptom for identifying urinary schistosomiasis. In this study, it was the most highlighted symptom of schistosomiasis. Awareness level of urinary, intestinal, female genital and male genital schistosomiasis were respectively low in this study. This reflects the general trend where more people are aware of urinary schistosomiasis than FGS (1,22,31). This trend could also arise from a lot of awareness creation and community efforts towards eliminating urinary schistosomiasis compared to FGS which, until recently, was not talked about much (36). Relatively low FGS awareness among health workers has been reported in studies in Ghana and Tanzania (22,32).

In this study, we assessed the association between FGS and schistosomiasis knowledge levels and various demographic factors, including job title, educational level, length of service, and gender. Only length of service showed a statistically significant association with FGS knowledge level (p = 0.003). This suggests that healthcare workers with longer service durations may have encountered FGS cases more frequently, even if they had previously overlooked them. Some gynecological clinical staff recalled instances of women from riparian communities presenting with persistent vaginal infections that did not resolve with standard treatment, a realisation that emerged after the questionnaire was administered and informal education on FGS was provided. For schistosomiasis knowledge levels, no demographic factor showed a significant association. This highlights a general gap in knowledge, reinforcing the need for continuous professional development programs to ensure healthcare workers remain updated on neglected tropical diseases. Poor awareness of FGS could contribute to misdiagnoses, affecting women’s reproductive health, increasing financial and mental health burdens, and ultimately hindering progress toward the WHO’s 2030 schistosomiasis elimination goal.

FGS has been implicated in the transmission of HIV (14,27) and HPV (37) among women in schistosomiasis-endemic regions. Knowledge of this would help health workers in considering FGS when women present with genital infections. Proper diagnosis and treatment would help resolve genital lesions and reduce the risk of contracting HIV. We therefore collated data on the awareness of FGS being implicated in the contraction of HIV and HPV. The results showed that out of the 68 people who answered these questions, majority (51) were aware of FGS playing a role in the contraction of HIV as compared to HPV (37). This disparity in awareness may stem from the greater emphasis in public health campaigns and literature on the link between HIV and other health conditions, including FGS, compared to HPV. HIV’s widespread attention as a global health issue likely contributes to higher recognition of its associations among healthcare workers as compared to HPV. Healthcare workers often mistake the symptoms and clinical signs of FGS for those of STIs due to their similarities (38) however, FGS is rarely included in their differential diagnoses or treatment plans unless the patient fails to respond to the initial therapy. Likewise, in a study conducted by Kukula et al. (22), adolescent girls exhibiting symptoms of FGS were often referred for treatment of STIs. This also presents another avenue for education to help reduce the risk of transmission in these areas.

After acquiring knowledge on FGS, health workers need resources to help diagnose and successfully treat. Colposcopy is one of the diagnostic methods for the diagnosis of FGS. It helps to visualise lesions in the vagina and then compare with the WHO FGS atlas as a guide (29). All facilities that took part in this study did not have a colposcope. This finding was the case in Wambui et al and Mazigo et al’s study (1,32) and other studies in sub-Saharan Africa (2,16). The lack of such an important gadget impedes diagnosis even if FGS is considered in differential diagnosis. The reason for not having this gadget by these facilities could be attributed to its high cost. An alternative diagnostic method will be PCR of cervicovaginal samples in the laboratory, but only tertiary and some secondary health facilities have such sophisticated equipment which is also expensive. Availability of praziquantel was also rare in the various facilities just like in studies conducted by Christinet et al., (2) Engels et al., (16) and Mazigo et al., (32). The head of one of the health facilities intimated that two pregnant women diagnosed with urinary schistosomiasis were once left to their fate due to lack of medication even at the district hospital.

We also assessed the human resources regarding the management of FGS. Staff who directly managed clients answered questions on how FGS is treated. Most of them got it right by choosing praziquantel, but others did not know, and some also chose antibiotics and antifungals, which were wrong. This indicates that mismanagement, whether due to a lack of knowledge about the appropriate treatment or the prescription of antibiotics or antifungals for misdiagnosed FGS cases, could place a significant burden on both patients and the healthcare system. Additional questions related to client care explored, whether health workers considered and differentiated FGS from sexually transmitted infections (STIs), STDs and other vaginal infections when diagnosing and managing female clients with such symptoms. The majority of respondents said “No” and some abstained from answering, indicating they did not. Just a few considered FGS during the diagnosis of genital issues. This implies that a lot of such cases will be missed and/or misdiagnosed, thereby causing future complications for these women. Regarding education on FGS, 92.83% of respondents have never had any form of education on the subject matter as was the case in a study conducted by Azanu et al., (35). FGS is a relatively recent focus of attention, even among clinicians, and is not yet included in Ghana’s educational institutions’ curriculum. During the FGS Accelerated Scale Together (FAST) package study, an in-person Continuous Professional Development (CPD) programme for Subject Matter Experts (SME) for the Ghana College of Medicine and Physicians, nursing training colleges and some faculties from some universities with the intention that FGS will be introduced into their curriculum (39). As a result, healthcare workers can only learn about it through specialised workshops and research articles. In contrast, schistosomiasis is covered in the curricula of most senior high schools and healthcare training institutions (35) in Ghana.

The perception of healthcare workers regarding FGS and schistosomiasis reveals notable trends in understanding and attitudes across various job roles. While most healthcare staff do not view clients with genital infections as promiscuous, some stigma persists, which underscores the need for training on non-judgmental, compassionate patient care. Furthermore, a strong majority of healthcare workers acknowledge both schistosomiasis as a public health concern and FGS as a significant threat to women’s reproductive health. This shared awareness likely reflects the widespread recognition of schistosomiasis’s impact on public health and underscores the urgent need for targeted interventions. Importantly, 231 respondents also see the need for awareness creation and education on FGS, suggesting a high level of commitment within the healthcare community to support advocacy and outreach efforts. Together, these insights point toward a robust foundation for enhancing FGS and schistosomiasis education and implementing stigma-reducing strategies within clinical practice.

## Conclusion

The study revealed a significant knowledge gap on schistosomiasis and female genital schistosomiasis among healthcare professionals in the Central Gonja District. This raises concerns about potential missed or misdiagnosed cases being classified as sexually transmitted infections. Addressing this gap is crucial, as it highlights the need for targeted capacity-building initiatives, such as workshops, in-service training programs, and clinical conferences, particularly for frontline healthcare staff involved in patient care. To enhance awareness and accurate diagnosis of FGS, it is recommended that future research focuses on intervention-based training programs and subsequent knowledge assessments. Such studies could also investigate the prevalence of FGS among genital infections reported at various health facilities, providing valuable data to guide public health interventions and improve patient outcomes.

## Data Availability

All relevant data are within the manuscript and its Supporting Information files.

## Acknowledgments

The authors sincerely thank all the individuals who participated in this study and generously shared their valuable experiences, particularly the healthcare workers in the Central Gonja District. We also extend our deep appreciation to the District Health Director of Central Gonja for his support and for facilitating engagement with facility heads across the district, which was instrumental to the success of this study.

## Author Contributions

**Conceptualisation:** Courage Gbeze, Akosua Bonsu Karikari, Nana Boakye Alahaman

**Data curation:** Courage Gbeze

**Formal analysis:** Courage Gbeze

**Methodology:** Courage Gbeze, Nana Boakye Alahaman

**Supervision:** Akosua Bonsu Karikari, Samuel Amoako Asirifi, Nana Boakye Alahaman **Validation:** Akosua Bonsu Karikari, Gloria Ivy Mensah, Kennedy Kwasi Addo **Writing – original draft:** Courage Gbeze

**Writing – review & editing:** Courage Gbeze, Akosua Bonsu Karikari, Nana Boakye Alahaman, Samuel Amoako Asirifi, Seth Christopher Yaw Appiah, Clémence Essé-Diby, Gloria Ivy Mensah, Kennedy Kwasi Addo

